# Metabolites from blood and formalin-fixed, paraffin-embedded tissue from participants with low- and high-grade prostate cancer: a pilot study

**DOI:** 10.64898/2026.03.12.26348192

**Authors:** Rebecca E. Graff, Henrik L. Bengtsson, Jung H. Suh, Adam B. Olshen, Elizabeth Y. Wang, Ryan M. Allen, Erin L. Van Blarigan, Stacey A. Kenfield, Janet E. Cowan, Peter R. Carroll, Jeff Simko, June M. Chan

## Abstract

**Background:** Identifying metabolites associated with prostate cancer (PC) aggressiveness may elucidate mechanisms underlying disease severity. Doing so for plasma and formalin-fixed, paraffin-embedded (FFPE) tissue could accelerate discovery. In this cross-sectional pilot study, we generated hypotheses for further exploration by assessing associations between plasma metabolites and Gleason score in individuals with PC and evaluating correlations between plasma and FFPE metabolite levels.

**Methods:** We examined plasma and FFPE samples from 10 individuals with Gleason score 7 (six 3+4, four 4+3) and nine individuals with Gleason score 9 (six 4+5, three 5+4) tumors from a convenience sample of 19 men with PC. We measured the relative abundance of polar metabolites at the time of radical prostatectomy. We used linear models of log_2_ fold changes to examine plasma metabolite levels relative to pathologic tumor grade. Relationships among metabolite levels measured in plasma and FFPE tumor tissue within individuals across metabolites were examined using Pearson correlations.

**Results:** Among 18 plasma metabolites selected *a priori* because of prior associations with PC aggressiveness, serine (*p*=0.0051) and ornithine (*p*=0.036) levels were higher in individuals with Gleason 9 than Gleason 7 PC. After multiple testing correction, however, no associations were statistically significant. The median correlation between levels in plasma and FFPE tumor tissue was 0.45 (range: 0.40-0.53) for the 94 metabolites measured in both biospecimens.

**Conclusions:** Plasma serine and ornithine demonstrated the largest differences between individuals with Gleason 7 and Gleason 9 PC. Metabolite levels in FFPE prostate tissue samples were moderately correlated with plasma levels. Future studies in larger samples are needed to further explore the hypotheses generated by this study.

## INTRODUCTION

Recent molecular and imaging tests have refined management of prostate cancer (PC),^1^ but it remains challenging to distinguish low-risk versus lethal disease at the time of diagnosis. Gleason score is among the strongest predictors of disease progression,^2^ but its utility in localized disease can be limited; men who undergo active surveillance are monitored with serial biopsies that may miss high grade foci of PC.^3–5^ A non-invasive method to detect high-grade tumors could improve disease monitoring and inform treatment decisions.

One such method that may prove fruitful is the evaluation of circulating metabolites. Metabolomic data have the potential to enhance the recognition of aggressive PC before it presents clinically. Emerging data have highlighted a number of intriguing metabolites and pathways in relation to PC aggressiveness.^6–9^ These studies have done so using a variety of biospecimens, including blood, urine, and frozen or fresh prostate tissue. Few studies have evaluated metabolites in formalin-fixed, paraffin-embedded (FFPE) prostate tumor samples,^10–12^ even though such specimens are routinely collected during medical care for PC and stable over time. Were it possible to accurately assay FFPE tissue, then many more studies would be able to evaluate the metabolomic basis of PC. Further, if circulating metabolite levels were reflective of the aggressiveness of the tissue metabolomic domain, then they could be leveraged to identify lethal disease early, when interventions are most effective.

We conducted this pilot study of PC metabolomics with two goals in mind: 1) to generate hypotheses for plasma metabolites associated with Gleason score at the time of radical prostatectomy (RP); and 2) to ascertain correlations between plasma, benign tissue, and tumor tissue metabolite levels both within individuals (across metabolites) and across individuals (for each metabolite).

## METHODS

### Study Population

The Department of Urology at the University of California, San Francisco (UCSF) maintains the Urologic Outcomes Database (UODB),^13^ which houses information on clinicopathologic, treatment, and outcomes data for individuals with PC undergoing active surveillance or RP. It additionally connects to a biobank that preserves biological specimens from urologic oncology patients. This pilot leveraged a convenience sample of 19 men treated with RP who contributed matched plasma and both benign and tumor FFPE prostate samples. An additional five men contributed matched benign and tumor FFPE prostate samples only. Plasma was extracted from banked fasting blood samples donated immediately prior to RP. Gleason scores of PC samples were determined by pathologic review of RP tissue.

All participants consented to the collection of samples for research purposes. The study protocol was approved by the institutional review board at UCSF.

### Metabolomic Profiling and Processing

Polar metabolomic analyses of all plasma and FFPE samples were performed with liquid chromatography with tandem mass spectrometry (LC-MS/MS), as has been described in prior publications.^10,14–16^ Metabolites with signal-to-noise ratio <5 or missing in ≥40% of the samples were omitted, after which peak intensities were normalized to metabolite median and centered. Specifically, the 5%-trimmed sample mean was calculated on the logarithmic scale for each participant and then transformed back to the original scale. The intensities were subsequently rescaled such that all participants had the same target average, which was arbitrarily defined as the median signal of the first participant.

### Statistical Analysis

To limit issues of multiple testing, we restricted analyses of plasma metabolite levels and Gleason score to 18 metabolites with prior evidence of a relationship with PC aggressiveness.^15,17–25^ To compare metabolite levels between individuals with Gleason score 9 versus 7 PC, we implemented linear models using the limma R package.^26^ We specifically utilized the standard limma model for differential expression that included an intercept and a term for Gleason score. The moderated t-test was then used to identify differences in metabolite values between individuals with Gleason scores of 9 versus 7. In sensitivity analyses, we compared individuals with 3+4 tumors to participants with 4+3 or 9 tumors.

Relationships among metabolite levels measured in plasma, benign prostate tissue, and tumor prostate tissue were examined using Pearson correlations, both within individuals across metabolites and across individuals for each metabolite.

All statistical analyses were carried out in R version 4.3.0.

## RESULTS

Among the 19 participants with available plasma, 10 had an RP Gleason score of 7 (6 with 3+4, 4 with 4+3) and nine had an RP Gleason score of 9 (6 with 4+5, 3 with 5+4) (**Table 1**). The median age of participants with available plasma at RP was 64 years (interquartile range: 11.5 years).

**Table 1.** Characteristics of 19 participants with plasma and FFPE tissue and five participants with FFPE tissue only.

| ID | Gleason Score | Age at RP | Tumor Tissue Site | Tumor % | Normal Tissue Site | Normal % |
| --- | --- | --- | --- | --- | --- | --- |
| **Plasma and FFPE** |  |  |  |  |  |  |
| 001 | 4+5 | 70-74 | Bilateral posterior mid | 90% | Left posterolateral mid | 10% |
| 002 | 5+4 | 70-74 | Right posterolateral mid | 95% | Left lateral mid | 10% |
| 003 | 4+3 | 70-74 | Right posterior mid | 80% | Left anterior mid | 20% |
| 004 | 5+4 | 60-64 | Bilateral posterior mid | 80% | Right anterior mid | 20% |
| 005 | 4+5 | 65-69 | Bilateral posterior apex | 70% | Left lateral mid | 10% |
| 006 | 4+3 | 55-59 | Bilateral posterior mid | 70% | Right anterior mid | 20% |
| 007 | 4+5 | 50-54 | Bilateral posterior apex | 90% | Right anterior apex | 20% |
| 008 | 3+4 | 45-49 | Bilateral posterior apex | 70% | Left anterior base | 40% |
| 009 | 4+3 | 65-69 | Bilateral posterior apex | 90% | Left lateral mid | 20% |
| 010 | 4+5 | 60-64 | Left lateral mid | 95% | Right lateral mid | 30% |
| 011 | 3+4 | 55-59 | Right posterior apex | 90% | Left lateral base | 30% |
| 012 | 3+4 | 55-59 | Left anterior mid | 90% | Right anterior base | 20% |
| 013 | 5+4 | 60-64 | Left lateral mid | 90% | Left posterior mid | 40% |
| 014 | 3+4 | 70-74 | Left posterolateral mid | 80% | Right posterolateral base | 30% |
| 015 | 4+3 | 65-69 | Left lateral apex | 90% | Right posterolateral mid | 40% |
| 016 | 4+5 | 60-64 | Left anterior apex | 95% | Bilateral posterior mid | 60% |
| 017 | 4+5 | 50-54 | Left posterolateral base | 80% | Left posterior mid | 50% |
| 018 | 3+4 | 50-54 | Left posterolateral mid | 90% | Right lateral mid | 30% |
| 019 | 3+4 | 65-69 | Bilateral central apex | 70% | Right posterolateral base | 30% |
| **FFPE Only** |  |  |  |  |  |  |
| 020 | 5+4 | 70-74 | Right lateral base | 70% | Right posterolateral base | 30% |
| 021 | 3+4 | 65-69 | Right lateral & anterior mid | 70% | Left posterior mid | 40% |
| 022 | 4+5 | 75-79 | Left lateral apex | 95% | Right lateral apex | 30% |
| 023 | 4+3 | 70-74 | Right lateral posterior mid | 90% | Bilateral anterior base | 30% |
| 024 | 3+4 | 70-74 | Posterior mid | 80% | Left anterior apex | 40% |

### Associations Between Plasma Metabolites and Gleason Score

**Figure 1** depicts a heatmap and hierarchical clustering of the 18 metabolites of interest according to Gleason score 7 versus 9 disease. No clear patterns emerged, nor were any of the metabolites statistically significantly associated with Gleason score following correction for multiple testing using the Benjamini-Hochberg method (**Table 2**). There were, however, two metabolites that were nominally associated with Gleason score based on linear models; higher levels of plasma serine (*p*=0.0051) and ornithine (*p*=0.036) were associated with Gleason 9 disease. Results for the remaining 98 metabolites successfully measured in plasma are presented in **Supplementary Table 1**). An additional three metabolites (2-aminooctanoic acid, acetyl-lysine, and sedoheptulose-1,7,-bisphosphate) demonstrated nominal evidence of an association.

**Figure 1.**
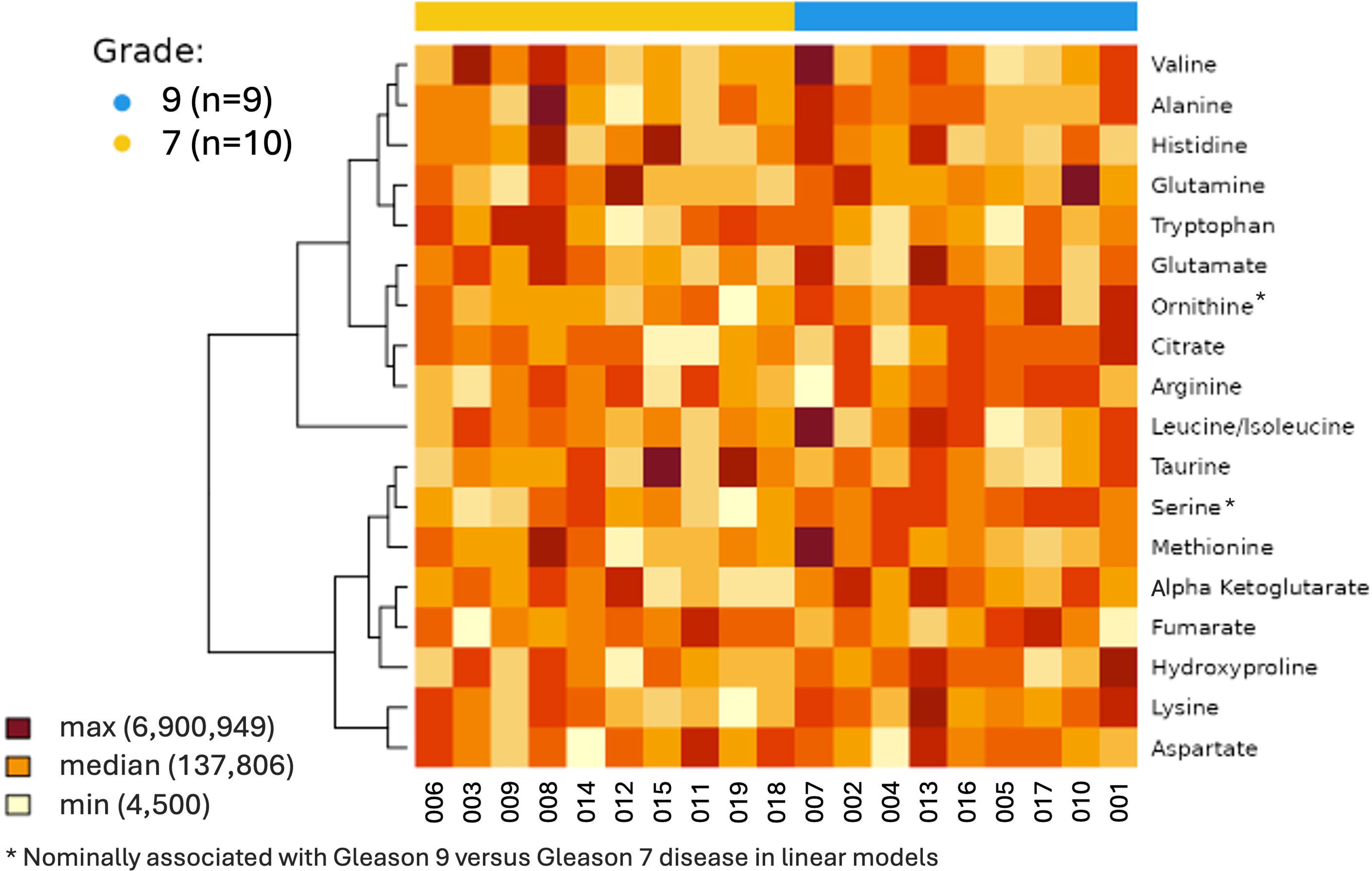
Heatmap and hierarchical cluster of log_2_ levels of 18 plasma metabolites of interest annotated by Gleason 7 (n=10) versus 9 (n=9) prostate cancer in 19 total participants. Rows were centered and scaled. Distances were computed using Euclidean distance, and hierarchical clustering was performed using the complete linkage method. The max, median, and min values are specified across all 18 metabolites on the original (i.e., not log_2_) scale.

**Table 2.** Associations between 18 plasma metabolites of interest and Gleason 7 (n=10) versus 9 (n=9) prostate cancer in 19 total participants.

| Metabolite | Gleason 7<br>Average Level | Gleason 9<br>Average Level | Fold Change<br>(95% CI) | p-value<sup>a</sup> | Adjusted<br>p-value<sup>a,b</sup> |
| --- | --- | --- | --- | --- | --- |
| Serine | 53268 | 70419 | 1.32 (1.10, 1.59) | 0.0051 | 0.092 |
| Ornithine | 162499 | 200814 | 1.24 (1.02, 1.50) | 0.036 | 0.32 |
| Lysine | 11649 | 13442 | 1.15 (0.96, 1.38) | 0.12 | 0.64 |
| Hydroxyproline | 24753 | 35304 | 1.43 (0.88, 2.31) | 0.14 | 0.64 |
| Tryptophan | 682931 | 596057 | 0.87 (0.69, 1.10) | 0.23 | 0.75 |
| Alpha Ketoglutarate | 22915 | 25102 | 1.10 (0.93, 1.29) | 0.27 | 0.75 |
| Glutamine | 712263 | 793665 | 1.11 (0.90, 1.38) | 0.30 | 0.75 |
| Citrate | 149618 | 171013 | 1.14 (0.86, 1.52) | 0.34 | 0.75 |
| Alanine | 415662 | 470913 | 1.13 (0.85, 1.51) | 0.38 | 0.75 |
| Arginine | 119288 | 127362 | 1.07 (0.87, 1.32) | 0.52 | 0.88 |
| Methionine | 42639 | 46449 | 1.09 (0.80, 1.49) | 0.58 | 0.88 |
| Fumarate | 22730 | 21277 | 0.94 (0.73, 1.20) | 0.59 | 0.88 |
| Taurine | 69946 | 66825 | 0.96 (0.77, 1.19) | 0.67 | 0.92 |
| Leucine/Isoleucine | 3710957 | 3864606 | 1.04 (0.80, 1.35) | 0.75 | 0.94 |
| Aspartate | 7334 | 7163 | 0.98 (0.80, 1.20) | 0.81 | 0.94 |
| Valine | 537528 | 547569 | 1.02 (0.79, 1.32) | 0.88 | 0.94 |
| Glutamate | 198975 | 202847 | 1.02 (0.77, 1.34) | 0.89 | 0.94 |
| Histidine | 314217 | 312434 | 0.99 (0.84, 1.18) | 0.94 | 0.94 |

Abbreviations: CI – confidence interval
<sup>a</sup> From linear models of log<sub>2</sub> metabolite levels
<sup>b</sup> Adjusted according to the Benjamini-Hochberg procedure

Sensitivity analyses that compared levels of the 18 metabolites of interest between the six individuals with Gleason 3+4 PC and the 13 participants with higher grade disease were similarly not statistically significant (**Supplementary Figure 1**, **Supplementary Table 2**). Only ornithine (*p* = 0.040) was associated with higher grade disease at a nominal significance threshold. Among the remaining 98 metabolites measured in plasma, only N6-acetyl-L-lysine and acetyl-lysine demonstrated nominal evidence of an association.

### Correlations Among Plasma, Benign Tissue, and PC Tissue Metabolites

Among the 116 metabolites that were successfully measured in plasma, 94 (81%) were also measured in the FFPE samples. Within individuals, correlations were generally stronger between benign and tumor tissue levels than between levels of either tissue measurement and plasma (**Supplementary Table 4**). Among 24 individuals with metabolite levels in benign and tumor tissue, the median correlation was 0.92 (range: 0.68-0.97). Among 19 individuals with available tissue and plasma metabolite levels, the median correlation between benign tissue and plasma levels was 0.47 (range: 0.40-0.54). Between tumor tissue and plasma metabolite levels, the median correlation was 0.45 (range: 0.40-0.53).

In analyses of metabolite levels across individuals for each metabolite, correlations between benign and tumor tissue were positive for 72 out of 94 metabolites (77%) and generally moderate in magnitude (median: 0.17) (**Supplementary Table 5**). Correlations between tumor tissue and plasma were more variable and frequently inverse (median: −0.09), whereas correlations between benign tissue and plasma were generally weaker and closer to null (median: −0.01). The metabolites that were most strongly positively correlated between tumor and plasma levels were urea (r^2^: 0.61), methylnicotinamide (r^2^: 0.53), and hypoxanthine (r^2^: 0.52).

## DISCUSSION

In this pilot study of PC metabolomics, we found suggestive evidence that circulating serine and ornithine may be associated with tumor Gleason score. We also achieved metabolite measurements in FFPE, values of which were moderately correlated with circulating metabolites levels within individuals. Metabolite levels in benign versus tumor FFPE were more strongly correlated. Across individuals, correlations between tissue and plasma levels for individual metabolites were variable and often modest.

Though results from this small pilot study should be treated as exploratory, mechanistic evidence suggests that both serine and ornithine could contribute to PC aggressiveness. Serine synthesis provides intermediates to the Krebs cycle, which sustains cancer metabolism.^27^ In addition, cells lacking p53 – a tumor suppressor often mutated in PC – may respond to therapy by serine depletion;^27,28^ increased serine would presumably work antagonistically. Ornithine could plausibly promote PC aggressiveness via its role in the synthesis of polyamines, which are involved in tumor growth, invasion, and development of metastases.^29,30^ In addition, ornithine is central to the urea cycle, dysregulation of which has also been implicated in prostate and other cancers.^31,32^ Both serine and ornithine are components of the glycine, serine, and threonine metabolism pathway, which refuels one-carbon metabolism, the process that cycles carbon units from amino acids to produce proteins, lipids, and nucleic acids essential to cell growth and proliferation.^27,33^ This pathway also plays an important role in cellular redox balance, fuels heme biosynthesis, and sustains oxidative phosphorylation,^34^ thereby increasing the proliferation rate of cancer cells.^33,35,36^

Indeed, preliminary data from other studies point to aberrant mobilization of glycine, serine, and threonine metabolism in aggressive PC.^11,15,17–19,24,25,29,37–61^ With respect to Gleason score specifically, one prior study identified urinary upregulation of the pathway as a whole in individuals with more poorly differentiated tumors.^45^ In another study, higher serine levels in seminal plasma were associated with higher Gleason score.^38^ Though previous studies have not identified associations between ornithine and tumor grade, several have found positive relationships with other PC characteristics indicative of aggressiveness (e.g., development of biochemical recurrence or metastases).^17,29,49,58^ Our results are thus consistent with the existing literature and mechanistically plausible, though they should be interpreted cautiously given the small sample size and only nominally significant p-values. They should also be validated in other studies.

FFPE tissue specimens are not often leveraged for studies of metabolomics due to concerns about metabolite changes during preservation. Nevertheless, preliminary studies support the feasibility and utility of MS-based metabolic profiling of FFPE tissue for PC^10–12^ and other tumor types.^62–71^ Those comparing metabolite levels in fresh-frozen versus FFPE prostate tumor tissue indicate that the latter have reduced coverage but can still recapitulate key biologic states.^10,11^ Our study adds to the limited existing body of evidence that FFPE prostate tissue can yield useful biochemical information. Within individuals, metabolite profiles were highly concordant between benign and tumor FFPE tissue, suggesting preservation of biologically meaningful metabolic patterns despite fixation. Correlations between tissue and plasma metabolite levels were more modest, indicating that circulating metabolites only partially reflect tissue metabolic profiles. In analyses across individuals for individual metabolites, concordance between plasma and tissue levels was variable, with only a subset of metabolites demonstrating moderate cross-matrix correlations. Together, these findings indicate that FFPE-derived metabolite measurements preserve biologically meaningful metabolic relationships within prostate tissue and support the use of archived FFPE specimens for metabolomic investigations of tumor biology and clinical characteristics. Given that FFPE tissue is more commonly available than other tissue types among individuals with long-term follow-up, metabolomics research leveraging FFPE samples has the potential to accelerate the identification of biochemicals that indicate PC aggressiveness.

Because our study was intended to pilot metabolomic analysis of samples from the UCSF urologic oncology biobank, its sample size was limited for meaningful biological inference. We were unable to adjust for secondary variables that would have strengthened causal interpretation. We nonetheless demonstrated the utility of our plasma and FFPE samples for future metabolomics research.

In conclusion, our results suggest that serine and ornithine are compelling metabolites for subsequent interrogation in the context of PC aggressiveness. They also demonstrate that metabolites are detectable in FFPE tissue samples, which could prove useful for future discovery of PC biomarkers. Such studies should include larger sample sizes and implement comprehensive covariate adjustment to disentangle systemic metabolic influences from tumor-specific biology.

## Data Availability

All data produced in the present study are available upon reasonable request to the authors

