## Supplementary Figure for "Metabolites from blood and formalin-fixed, paraffin-embedded tissue from participants with low- and high-grade prostate cancer: a pilot study"

**Supplementary Figure 1**. Heatmap and hierarchical cluster of log_2_ levels of 18 plasma metabolites of interest annotated by Gleason 3+4 (n=6) versus >3+4 (n=13) prostate cancer in 19 total participants. Rows were centered and scaled. Distances were computed using Euclidean distance, and hierarchical clustering was performed using the complete linkage method. The max, median, and min values are specified across all 18 metabolites on the original (i.e., not log_2_) scale.

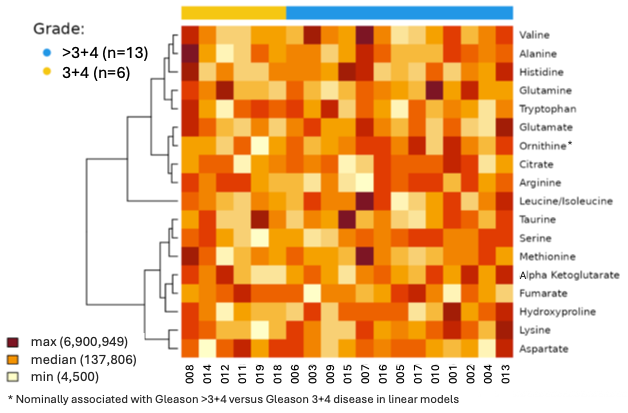
